# Efficacy of SARS-CoV-2 wastewater surveillance for detection of COVID-19 at a residential private college

**DOI:** 10.1101/2021.09.15.21263338

**Authors:** Michelle Landstrom, Evan Braun, Ellen Larson, Merrill Miller, Geoffrey H. Holm

**Affiliations:** Department of Biology, Colgate University, 13 Oak Dr., Hamilton, NY, 13346; Student Health Services, Colgate University, 13 Oak Dr., Hamilton, NY, 13346

**Keywords:** SARS-CoV-2, COVID-19, wastewater surveillance, sewershed, residential college

## Abstract

Many colleges and universities utilized wastewater surveillance testing for SARS-CoV-2 RNA as a tool to help monitor and mitigate the COVID-19 pandemic on campuses across the United States during the 2020-2021 academic year. We sought to assess the efficacy of one such program by analyzing wastewater RNA load data in relation to SARS-CoV-2 cases identified through individual surveillance testing. Almost 80% of the cases on campus were associated with positive wastewater tests, resulting in an overall positive predictive value of ∼79% (Chi square 48.1, Df = 1, *p* < 0.001). However, half of the positive wastewater samples occurred in the two weeks following the return of a student to the residence hall following isolation, and therefore were not useful in predicting new infections. When these samples were excluded, the positive predictive value of a positive wastewater sample was ∼54%. Overall, we conclude that the continued shedding of viral RNA by patients past the time of potential transmission confounds the identification of new cases using wastewater surveillance, and decreases its effectiveness in managing SARS-CoV-2 infections on a residential college campus.

## Introduction

The severe acute respiratory syndrome coronavirus 2 (SARS-CoV-2) pandemic clearly poses significant challenges for many economic sectors. Prior to the introduction of vaccines, and even post-introduction, many businesses and institutions that rely on in-person meetings struggled to identify best practices that could allow for continued operation. In some instances, especially during government-mandated “lockdowns,” institutions transitioned to remote operations. However, in many cases, remote operation was either not sustainable or failed to deliver satisfactory outcomes, leading these institutions to consider means for returning to in-person operation. These challenges continue to be felt acutely among institutions of higher education, particularly those with a sizeable residential component, as the high population density of dormitories, especially those with shared bathroom facilities, create conditions that can facilitate rapid transmission of respiratory pathogens.

As data emerged that SARS-CoV-2 can infect cells in the gastrointestinal tract (1, 2), and that SARS-CoV-2 RNA can be detected in patient stool (3, 4), including from patients prior to the onset of clinical symptoms (5), many groups began investigating whether detection of SARS-CoV-2 RNA in wastewater could be used as a means of monitoring communities for emerging infections, or of directing public health responses. Epidemiological wastewater surveillance had previously been utilized for poliovirus, norovirus, and rotavirus (6–8), and gave strong indication that similar approaches could be beneficial, either to directly assess disease burden in a population, identify areas where the SARS-CoV-2 was newly introduced, or monitor changes in transmission status over time and thereby assess the effects of community interventions (9). The utility of these approaches became even more evident as data emerged correlating levels of SARS-CoV-2 RNA in wastewater with number of cases in the sewershed (10–12), with wastewater RNA levels increasing several days prior to an increase in positive test results indicative of SARS-CoV-2 infections (13–15).

Based on these reports, in the lead up to the Fall 2020 academic semester, many colleges and universities in the U.S. sought to implement wastewater surveillance as a tool to manage the pandemic, either as a means to detect the presence of cases within the campus population, to monitor infection levels on campus, or to target particular residence halls for individual or pooled patient testing to directly identify infected individuals (16–21). Since essentially all of these systems were implemented *de novo*, there was a lot of variability in how they were set up and used. Some institutions monitored wastewater via campus or municipal wastewater treatment plants, whereas others implemented systems to capture samples from individual residence halls or sewer lines. Some institutions collected samples that were then processed by commercial laboratories, whereas others handled their samples in house. Among the strategies communicated by various institutions, there was considerable variation in the timing and frequency of collection, and in the number of campus locations that were sampled. Given this variability, it is important to assess the efficacy of the various approaches, to identify best practices and help inform continued development and implementation of wastewater surveillance strategies as a pandemic response (19).

In the summer of 2020, we developed and implemented a wastewater surveillance system as one piece of a comprehensive plan to facilitate the return of students to campus residences at Colgate University. Wastewater was assayed via 24-hour composite samples taken 2-3 times weekly at each of seven campus locations (sewer lines or sewage lift stations) and the samples were tested for the presence of SARS-CoV-2 RNA by quantitative reverse-transcriptase PCR (qRT-PCR). Simultaneously, students were regularly tested for SARS-CoV-2 infection by anterior nares swabs, using a commercial PCR-based testing provider. We obtained de-identified information on student positive cases, the Ct value of the positive tests, and the time of removal and return to the residence hall for isolation, and analyzed this data with respect to the detection of SARS-CoV-2 in the waste stream. In the fall and spring academic semesters, Colgate identified a total of 107 student cases of SARS-CoV-2 infection. Almost 80% of these cases were associated with positive wastewater tests, resulting in an overall positive predictive value of ∼79%. However, half of the positive wastewater samples occurred in the two weeks following the return of a student to the residence hall following isolation. When these samples were excluded, the positive predictive value of a positive wastewater sample was ∼54%. Overall, we conclude that the continued shedding of viral RNA by patients past the time of potential transmission confounds the identification of new cases using wastewater surveillance, and decreases its effectiveness in managing SARS-CoV-2 infections on a residential college campus.

## Materials and Methods

### Study site

Colgate University is a private residential undergraduate college with around 3,000 students, situated in the rural community of Hamilton, NY (population ∼3,800). For the 2020-2021 academic year, students had the choice of remote or in-person instruction, and over 2,600 students chose to be in residence, with 2,333 students in campus residence halls in the fall semester and 2280 students in campus residence halls in the spring semester (Table 1), and the rest in off-campus (private) housing in the village of Hamilton.

**Table 1.** Student population, cases, and positive wastewater samples at Colgate University, Fall 2020-Spring 2021.

| Sampling Locations | Fall Population | Spring Population | Fall Cases | Spring Cases | Fall Positive WW samples | Spring Positive WW samples |
| --- | --- | --- | --- | --- | --- | --- |
| GH/BAE | 336 | 336 | 4 | 7 | 4 | 7 |
| SWP | 289 | 269 | 3 | 5 | 3 | 5 |
| CD | 338 | 330 | 6 | 13 | 0 | 13 |
| BC | 370 | 365 | 5 | 6 | 2 | 2 |
| PL | 162 | 160 | 7 | 5 | 2 | 12 |
| TH | 219 | 212 | 6 | 12 | 1 | 9 |
| **Total Student Population/Cases in Sampled Residences (percent of total)** | **1714 (73.5%)** | **1672 (73.3%)** | **31 (70.5%)** | **48 (76.2%)** |  |  |
| **Total Student Population/Cases in Non-Sampled Campus Residences** | **619 (26.5%)** | **608 (26.7%)** | **13 (29.5%)** | **15 (23.8%)** |  |  |
| **Total** | **2333** | **2280** | **44** | **63** |  |  |

### Wastewater sample collection

Wastewater samples were collected at seven locations on the Colgate University campus (Table 1; Fig. 1) using three ISCO GLS compact composite samplers (Teledyne ISCO, Lincoln, NE). Five of the seven locations (GH, SPW, BAE, CD, BC) were in sewer access holes, while two were at lift stations (PL, TH). A 24 h composite sample (50 ml every 20 min) was collected at around 10 AM each weekday, and the samplers were rotated among the collection points at that time, resulting in each collection point being sampled an average of 2.1 times per week. Three of the access points (GH, BC, and TH) required external ground-level storage of the sampler; at these locations, a single draw was obtained during the times when the ambient air temperature was too low to permit composite sampling (intermittently between Jan. and Mar.). Composite samples were mixed by agitation and ∼250 ml of the total volume of each sample was collected and returned to the lab, where it was stored at 4°C until processing. The interior tubing of the samplers was rinsed with 10% bleach followed by distilled water after each sample collection. Dedicated collection hoses and filters remained at each location and were rinsed with distilled water after each collection and with 10% bleach on a regular basis. Samples were also collected from the Village of Hamilton wastewater treatment plant, though these samples were excluded from this analysis since infection data from the entire population serviced by this facility (which includes Colgate University and Colgate’s isolation space) was not available.

**Figure 1.**
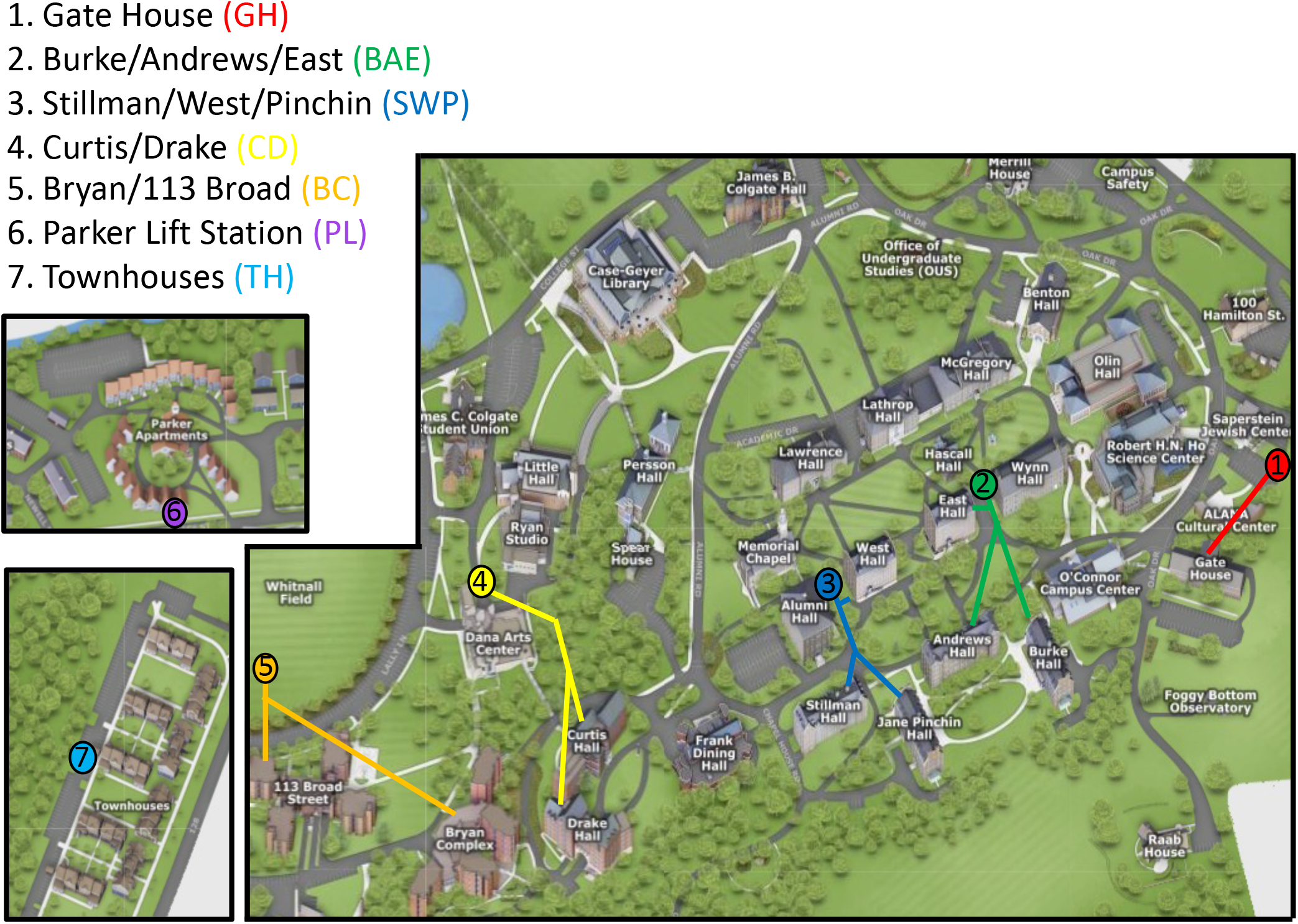
Location of wastewater sample collection points on the Colgate University campus. Locations 1-5 were at sewer access holes, while locations 6-7 were at wastewater lift stations. Colored lines indicate approximate location of sewage drains from each residence.

### Ultracentrifugation and RNA extraction

RNA was extracted following the method of Wilder et al. 2021 (22). All steps prior to RNA extraction were conducted in BSL2 conditions, as approved by the Colgate University Institutional Biosafety Committee. Briefly, ∼26 ml of composite sample was layered over 12 ml of sucrose cushion (50% sucrose in 20 mM Tris-HCl (pH 7.0), 100 mM NaCl, and 2 mM EDTA) in a 25×89 mm ultracentrifuge tube (Beckman Coulter, Brea, CA). The samples were then centrifuged in a SW28 Ti rotor (Beckman Coulter) at 150,000 x *g* in a L7-55 ultracentrifuge (Beckman) for 45 m at 4°C. The supernatant was decanted and the pellets were resuspended in 200 µl of PBS and transferred to a 1.7 ml microcentrifuge tube. RNA was extracted from the sample either using an AllPrep PowerViral DNA/RNA kit (Qiagen Biosciences) according to manufacturer’s protocol (with the omission of the optional bead beating step), or using Trizol (1 ml; Invitrogen/ThermoFisher) according to manufacturer’s protocol. Both methods yielded equivalent RNA isolation based on pilot runs using duplicate samples. RNA was eluted in 50 µl of RNase-free water and stored (when necessary prior to analysis) at −80°C. Samples (5 µl) were treated with DNase I (Invitrogen/ThermoFisher) according to the manufacturer’s protocol. DNase-treated samples were then directly used for quantitative reverse transcriptase PCR (qRT-PCR).

### Quantification of relative wastewater SARS-CoV-2 RNA levels

Levels of SARS-CoV-2 RNA in undiluted samples were detected by one-step multiplex TaqMan qRT-PCR, using TaqPath 1-step Multiplex Master Mix (ThermoFisher) according to manufacturer’s protocol. SARS-CoV-2 RNA was detected using the 2019-nCoV CDC RUO kit (Integrated DNA Technologies; IDT), containing primers amplifying two regions in the SARS-CoV-2 nucleocapsid (N) gene (N1 and N2). A control plasmid containing the complete nucleocapsid gene from SARS-CoV-2 (IDT), as well as a synthetic SARS-CoV-2 RNA (Applied Biological Materials, Richmond, BC, Canada), were used as positive controls for isolation procedures and for qRT-PCR reactions. To normalize for differences in biological material in each sample, extracted RNA was also probed using TaqMan primers specific for human RNAse P (2019-nCoV CDC RUO primers and probe kit; IDT), and for the crAssphage bacteriophage (22). Initial samples showed a high degree of correlation between RNAse P Ct values and crAssphage Ct values, although crAssphage was not as reliably detected in all samples (not shown). Therefore, RNAse P Ct values were used to normalize SARS-CoV-2 N gene multiplex Ct values, using the ΔΔCt method, to obtain a relative value for SARS-CoV-2 RNA for each sample, which was reported to the Colgate University Health Analytics Team.

### Individual SARS-CoV-2 surveillance testing

For both academic semesters, all Colgate University students were required to submit negative results from a home PCR-based SARS-CoV-2 test taken ∼7 days prior to arrival at campus. Students, along with all employees, were then tested using a PCR-based test at the time of student arrival to campus. All students then entered a mandatory 14-day quarantine period. Students were tested a second time ∼7-9 days post-arrival and were not released from quarantine until the results from the second test had been returned. Following quarantine, the whole campus population (students and employees) were randomly selected for surveillance testing at a rate of 6% of each residence hall/area or campus employee population per week (fall semester), or scheduled for surveillance testing once every 2 weeks (spring semester). In the fall semester, ∼50-70 additional students were selected each week for targeted surveillance testing based on wastewater SARS-CoV-2 results, identified positive cases within specific residence halls, or other events indicating increased risk of transmission (bringing the total percentage of the student population tested up to ∼10% per week). Targeted surveillance testing was not widely performed during the spring semester, given the increased frequency of baseline surveillance testing. Patient samples were collected from the anterior nares by Colgate University Student Health Services and volunteers, and were shipped to Aegis Biosciences (Nashville, TN) for PCR-based analysis. Ct values for positive tests were reported by Aegis to Colgate University Student Health Services. The average Ct value for two or three PCR reactions using individual primer pairs was calculated for each case and used for analysis. In both semesters, students with symptoms consistent with COVID-19 were diagnostically tested in-house using a rapid PCR-based assay (Cepheid Biosciences). In some cases, patient tests were performed at other local laboratories. Ct values were not available for these test results. Students that tested positive for SARS-CoV-2, and their close contacts, were identified upon receipt of test results (typically 48-72 h post-collection) and separately isolated or quarantined at a local facility (which was not subject to wastewater surveillance), or, if feasible, at their family home, until allowed to return to their residence hall as established by Madison County Board of Health and New York State guidelines.

### Data analysis

Data on positive SARS-CoV-2 test results, their Ct values (if available), and the start and return dates of isolation, devoid of patient-identifying information were obtained from Colgate University Student Health Services. Data on de-identified total residence hall occupancies were obtained from the Office of Institutional Planning and Research. Due to low case numbers in one residence (GH), data from this residence and waste stream was combined with that of BAE to eliminate potentially identifying information. Additionally, specific dates have not been included in the manuscript to also eliminate potentially identifying information. This project was ruled as exempt from the review process by the Colgate University Institutional Review Board. Data was analyzed using GraphPad Prism statistical software package as indicated. For temporal analysis, positive wastewater samples were categorized according to their temporal relationship with positive student case(s), including those that occurred: i) within seven days prior to detection of a positive case; ii) between the date of a positive patient sample and when the student was moved to the isolation facility; iii) between the date a student was moved to the isolation facility and when they returned to the residence; iv) within 5 days of the student’s return from isolation; v) within 6-14 days of the student’s return from isolation; and vi) those that did not correspond to any identified case within those temporal parameters.

## Results

During the spring and summer of 2020, Colgate University engaged in a strategic planning process to determine how best to facilitate the return of students to campus for the 2020-2021 academic year, given the challenges associated with the SARS-CoV-2/COVID-19 pandemic. At that time, data was emerging regarding the efficacy of detecting the presence of SARS-CoV-2 RNA in wastewater efflux via quantitative reverse transcriptase PCR (qRT-PCR) as a means of population surveillance. SARS-CoV-2 RNA can be detected in the wastewater stream as early as a week prior to identification of patients with COVID-19 based on screening of symptomatic individuals (13, 14), and based on this information, we decided to implement wastewater surveillance as one part of a comprehensive plan to facilitate the return of students to campus residences for the academic year. Colleagues at other local academic institutions were working on methods to quantify SARS-CoV-2 RNA in wastewater (22) and graciously assisted in developing protocols for wastewater surveillance at Colgate University.

The University purchased three compact composite wastewater samplers and we consulted with the Facilities Department to determine locations for sampling. We identified seven locations on campus that would allow as many student residences as possible to be sampled, while minimizing the overlap with effluent waste from academic and administrative buildings (Fig. 1). However, due to the integrated nature of some administrative spaces within residence halls, it was impossible to exclude these entirely at some of the sampling points. In total, the sampling locations captured waste from thirteen individual residence halls or residential complexes, which housed approximately 73% of Colgate students living in campus-owned residences (Table 1). The largest population sampled was a complex with 370 students, while the smallest was a residence with 73 students, with a mean of 242 residents sampled at each collection point. Due to containing potentially identifying information, data from the smallest residence (GH) was combined with that for BAE for analysis. For wastewater samples, the relative level of SARS-CoV-2 RNA was determined using the Ct value of multiplex qRT-PCR reactions targeting the SARS-CoV-2 genome, normalized to the amount of biological material in the sample, as determined by the Ct value of a qRT-PCR reaction to detect human RNase P mRNA.

Concurrently, Colgate University engaged in individual surveillance testing of students and employees, as outlined. During the fall semester, from the start of first year arrival in late August, 2020 until most students left campus in mid-November, PCR-based surveillance testing detected 44 cases of SARS-CoV-2 infection in the student population (Table 1; Fig. 2A). Of the fall cases, 36 of the 44 (81%) were detected during the two arrival testing periods, and many of these cases had Ct values of >35, suggesting low viral RNA levels. The mean Ct value of the positive tests in the fall was 34.54 (Fig. 3A). Thirty-one of the cases (70.5%) occurred in residences from which wastewater samples could be collected. In the spring semester, from arrival for most students starting in late January, 2021, through the end of the semester in early May, 63 cases of SARS-CoV-2 infection were detected (Table 1; Fig. 2B). Again, a large number of cases (28; 44%) were detected during the two arrival testing periods. The mean Ct value of the positive tests in the spring was 26.59, which was significantly lower (higher viral RNA levels) than in the fall (Fig. 3A; *p* < 0.0001). The percentage of cases that occurred in residences from which wastewater samples could be collected (48 of 63; 76.2%) was not significantly different than in the fall (*p* = 0.51 by Chi-square analysis).

**Figure 2.**
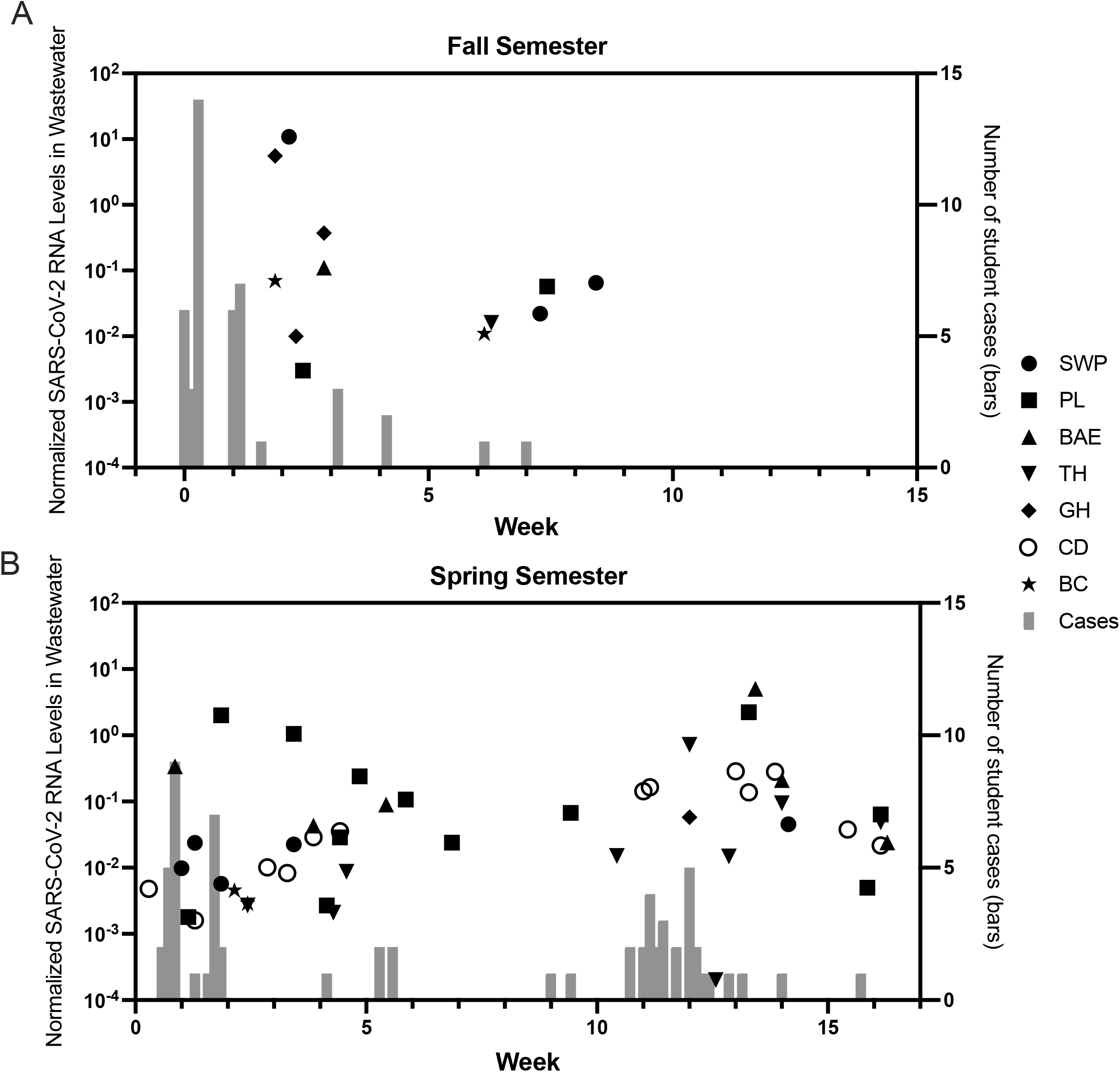
Aggregate wastewater and patient case data at Colgate University, August 2020-May 2021. Normalized SARS-CoV-2 RNA levels in wastewater (black symbols; left Y-axis) and number of positive student cases (gray bars; right Y-axis) are shown from the fall semester 2020 (A) and spring semester 2021 (B). Symbol shapes represent individual sampling locations and associated residence halls, as indicated in the legend.

**Figure 3.**
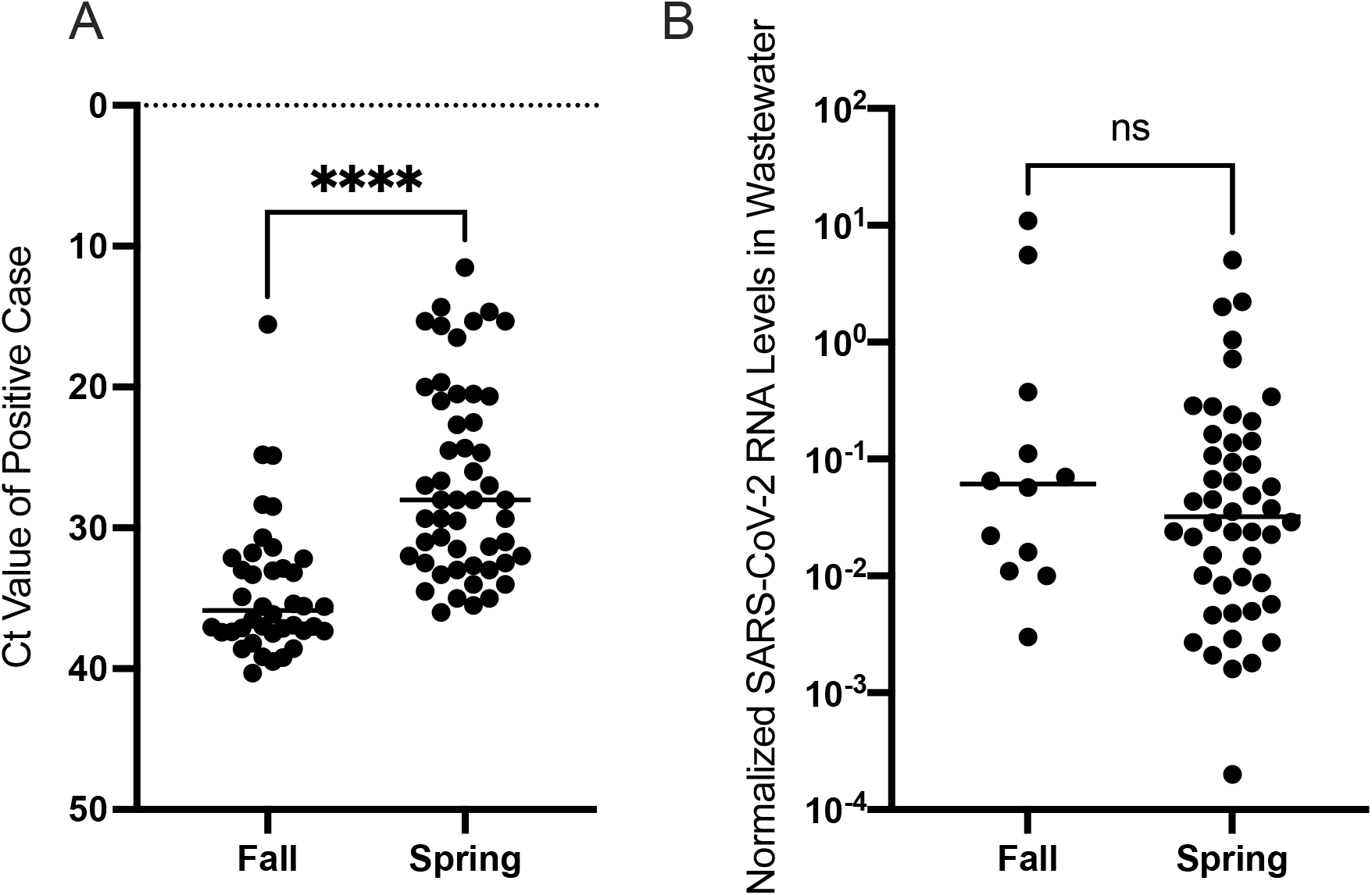
The relative level of SARS-CoV-2 RNA in positive SARS-CoV-2 clinical cases, but not the levels of wastewater RNA, were significantly higher in the spring semester. The data points represent the average Ct value from positive cases (A) and the normalized level of SARS-CoV-2 RNA levels in positive wastewater samples (B). ****, *p <* 0.0001 by Welch’s t test. ns, *p* > 0.05 of log transformed data by Welch’s t test.

In the fall, SARS-CoV-2 RNA was detected in 12 wastewater samples, from 6 out of the 7 collection points (Table 1; Fig. 2A). During the spring semester, 48 wastewater samples tested positive for SARS-CoV-2, from all 7 collection points (Table 1; Fig. 2B). The relative level of SARS-CoV-2 RNA detected in wastewater samples was not significantly different between the two semesters (Fig. 3B). We analyzed the data for each wastewater sample location (Fig. 4), plotting the relative wastewater SARS-CoV-2 levels (black circles; left Y-axis); as well as the Ct value of each positive patient sample (red circles; right Y-axis). We also plotted positive patient samples for which no Ct value was available (red stars), as well as the dates the positive cases were moved from the residence hall into the isolation facility (yellow squares) and the dates the cases moved back to the residence hall following isolation (green squares). From these data, we categorized each positive wastewater sample according to its temporal relationship with positive student case(s), as described.

**Figure 4.**
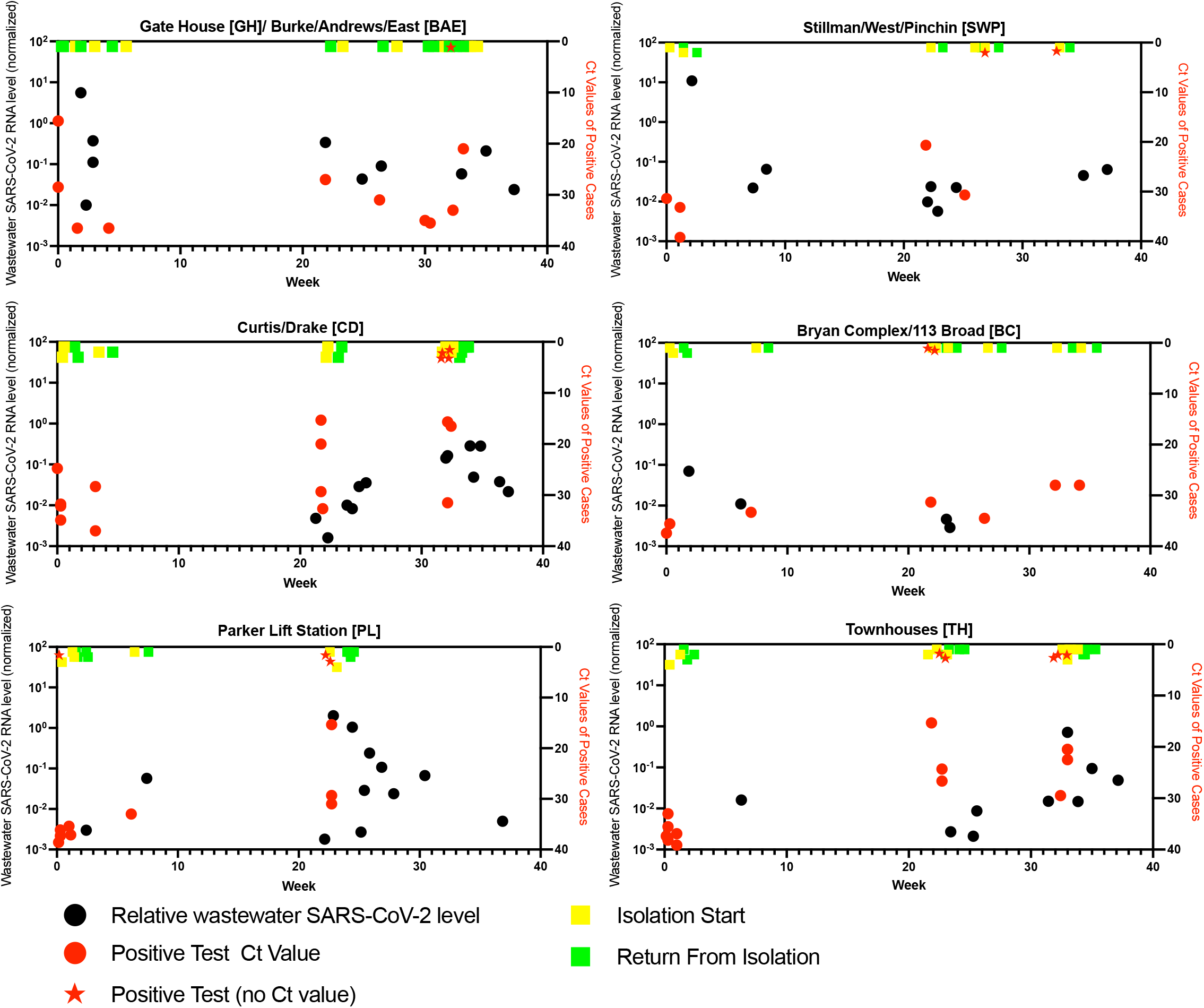
Aggregate wastewater and patient case data for sampled sewersheds, Colgate University, August 2020 -May 2021. Normalized SARS-CoV-2 RNA levels in wastewater (black circles; left Y-axis) and average Ct values of positive cases (red circles; right Y-axis) are shown from each wastewater sampling location. Red stars indicate positive student tests for which no Ct value was available. Yellow and green squares represent the time of isolation start (when individual student cases were removed from the indicated residences) and the time of return to the residence following isolation, respectively.

Overall, 14 of the positive samples (23.3%) were collected in the seven days prior to the identification of an infected individual in a residence that contributed to the sampled waste stream, or in the time between when patient specimen was collected and the time the individual moved to quarantine (Fig. 5A). Thirty of the positive samples (50%) were collected from locations in the two weeks following the return of an individual from isolation into that waste stream. Only two positive samples were collected in the period between when an infected individual was removed to the isolation facility and when they returned to the residence, which could result from lingering biological material in the waste stream. Twelve positive samples (20%) were not linked temporally with any student cases in that waste stream. There was no significant difference in the relative wastewater SARS-CoV-2 RNA levels in samples from these categories (Fig. 5B; *p* = 0.2 by Kruskal-Wallace test). We also analyzed the wastewater results according to the number of student cases identified in the residences contributing to the waste stream at the time of sample collection. Again, no significant differences were observed (Fig. 5C; *p* = 0.17 by Kruskal-Wallace test).

**Figure 5.**
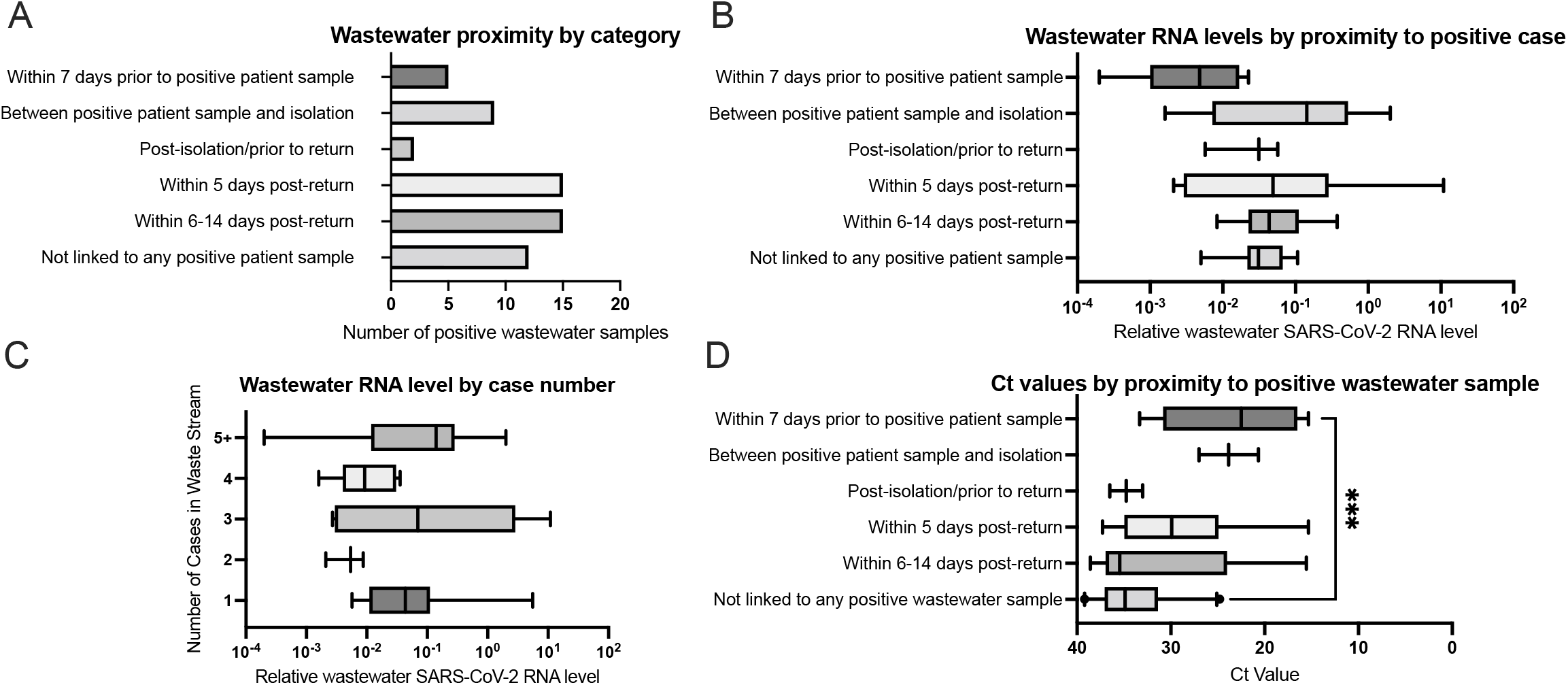
Temporal association of SARS-CoV-2 levels in wastewater RNA and Ct values of positive cases. Data from individual wastewater SARS-CoV-2 RNA samples and positive SARS-CoV-2 cases were classified according to the indicated temporal relationships. A. The number of positive wastewater samples according to temporal category, with respect to positive case in the residences contributing to the waste stream. B. The relative wastewater SARS-CoV-2 level according to temporal category, with respect to positive case(s) in residences contributing to the waste stream. C. The relative wastewater SARS-CoV-2 level according to the number of clinical cases identified in residences contributing to the waste stream at the time of collection. D. The relative Ct value of positive case(s) according to temporal category, with respect to positive detection(s) of SARS-CoV-2 in the waste stream. Data represent total number of samples (a) or a box and whisker plot indicating the 5th and 95th percentile (B,C). ***, *p* < 0.001 by Kruskal-Wallis test, with Dunn’s multiple comparison.

A similar analysis was conducted on the Ct values of the patient specimens based on the temporal proximity of positive wastewater samples (Fig. 5D). In this analysis, 21 of the identified student cases in sampled residences (21.5%) were not linked temporally with any positive wastewater sample. There was a significant difference in Ct values between categories, with significantly higher Ct values (lower viral RNA levels) found in patient samples that were not linked with any positive wastewater samples than in those found in patients in which a positive wastewater sample was detected in the seven days prior to specimen collection (*p* < 0.001 by Kruskal-Wallace test with Dunn’s multiple comparisons).

Examining the ability of a positive wastewater test to predict the presence of a case in a residence, we found that 46 positive wastewater samples were collected from residences in which a student case was identified, in the period from 7 days prior to the positive patient sample(s) being collected to 14 days after the student(s) returned to the residence following isolation. In contrast, 12 positive wastewater samples were collected from residences in which no student case was identified (i.e. false positives), resulting in an overall positive predictive value of ∼79% (Table 2; Chi square 48.1, Df = 1, *p* < 0.001). Out of 350 negative wastewater samples, 120 were collected in the period from 7 days prior to a positive case identification to 14 days after the student(s) returned to the residence (i.e. false negatives), resulting in an overall negative predictive value of ∼66%. From this data, the sensitivity of the wastewater testing was ∼28%, while the specificity was ∼95%. The fact that many of the positive samples occurred in the 14 day period following students returning to a residence following isolation was not unexpected, as SARS-CoV-2 RNA can be detected in patient stool for several weeks following infection (23). However, this could confound the use of wastewater samples to identify new cases in a residence hall. To assess the predictive capacity for an isolated positive wastewater sample to identify new cases in residence halls with no recent cases, we excluded all samples collected in the 14 days following the return of a student to a residence from the analysis. We identified 14 positive wastewater samples collected in the period 7 days prior to when a positive patient sample was collected, in contrast to 10 negative samples collected in the same period, giving a sensitivity of ∼58% (Table 3; Chi square 64.61, Df = 1, *p* < 0.001). The positive predictive value of these positive samples, based on the 12 false positive samples, was

**Table 2.**
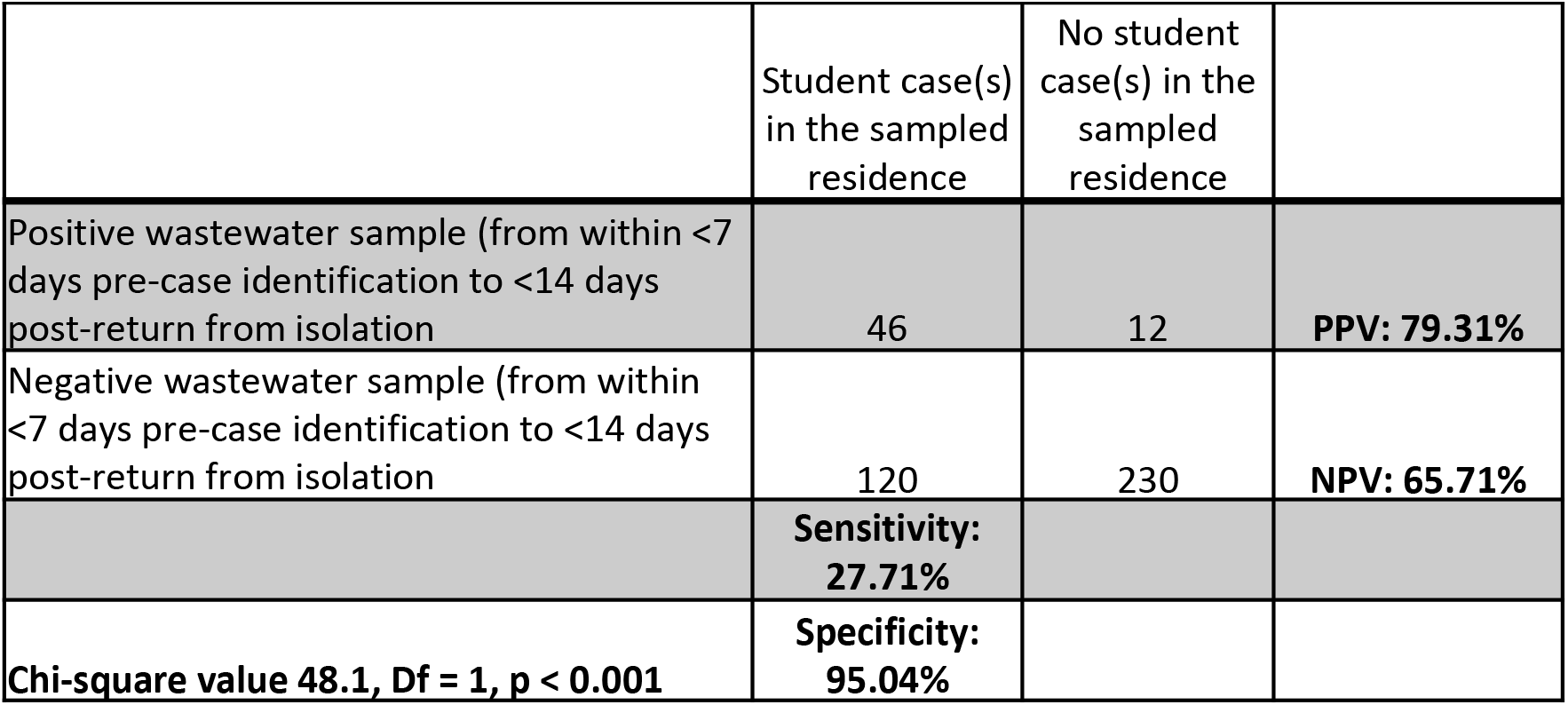
Performance of SARS-CoV-2 wastewater testing (within <7 days pre-case identification to 14 days post return from isolation) in predicting human cases at Colgate University, Fall 2020-Spring 2021

|  | Student case(s) in the sampled residence | No student case(s) in the sampled residence |  |
| --- | --- | --- | --- |
| Positive wastewater sample (from within <7 days pre-case identification to <14 days post-return from isolation) | 46 | 12 | **PPV: 79.31%** |
| Negative wastewater sample (from within <7 days pre-case identification to <14 days post-return from isolation) | 120 | 230 | **NPV: 65.71%** |
|  | **Sensitivity: 27.71%** |  |  |
| **Chi-square value 48.1, Df = 1, p < 0.001** | **Specificity: 95.04%** |  |  |

**Table 3.** Performance of SARS-CoV-2 wastewater testing (within <7 days pre-case identification) in predicting human cases at Colgate University, Fall 2020-Spring 2021

|  | Student case(s) in the sampled residence | No student case(s) in the sampled residence |  |
| --- | --- | --- | --- |
| Positive wastewater sample (from within <7 days pre-case identification) | 14 | 12 | **PPV: 53.85%** |
| Negative wastewater sample (from within <7 days pre-case identification) | 10 | 230 | **NPV: 95.83%** |
|  | **Sensitivity: 58.33%** |  |  |
| **Chi-square value 64.61, Df = 1, p < 0.001** | **Specificity: 95.04%** |  |  |

∼54%, while the negative predictive value was ∼96%.

Overall, this data suggests that wastewater testing deployed in this manner can be largely predictive of SARS-CoV-2 infections in college residence halls; however, the analysis of a positive result can be complicated in situations where infected students return to a residence following isolation, as SARS-CoV-2 RNA may continue to be detected in the waste stream for at least two weeks after return.

## Discussion

Municipal wastewater testing for SARS-CoV-2 RNA can provide a robust indicator of community transmission (11, 13, 14), and this information can be used to inform public health responses. Many colleges and universities implemented wastewater surveillance testing during the 2020-2021 academic year (19); however, the efficacy of these efforts in identifying infection clusters and leading to mitigation of transmission has not been fully investigated. In this study, we combine data on wastewater SARS-CoV-2 detection from discrete campus residences with information on student cases, including RNA levels and timing of exit and return to the residence following isolation, to provide insight into the efficacy of wastewater surveillance as a public health strategy at a residential undergraduate college. Overall, we found that while around a quarter of positive wastewater samples indicated the presence of SARS-CoV-2-infected individuals prior to their identification by individual surveillance testing, half of the positive wastewater samples occurred within 14 days after students returned to residence halls following isolation. The capacity for individuals to continue to shed SARS-CoV-2 RNA in stool past the point of likely transmission potentially limits the utility of wastewater surveillance in a college environment where students leave and return to residences following infection.

Our results are generally concordant with other reports of wastewater surveillance programs on college campuses. Our determined positive predictive value for isolated wastewater positive samples (excluding dormitories with students returning from isolation), of ∼54% is lower than some reported values, such as at the University of Arizona (82%; (21)), or at UC San Diego, where wastewater surveillance allowed for the early diagnosis of 85% of cases on campus (16). However, these programs operated on a very different scale, with, for example, the UCSD system employing 68 samplers with automated sample processing of daily samples. Similar to other institutions, we could identify single cases in a residence hall from wastewater samples (17, 18), although a large number of negative samples from residences with student cases decreased the overall sensitivity. Groups that sampled on a weekly basis concluded that that was too infrequent for effective identification of cases, and suggested that daily testing would be most appropriate (20). We found that our strategy, where most residences were sampled 2-3 times a week, was generally effective while not becoming impractical to manage for a small academic laboratory.

We observed significantly higher average Ct values (lower viral RNA levels) among cases in the fall semester than the spring semester (Fig. 3A). This is most likely a reflection of changes in the stringency of the cutoff for the identification of a positive case used by the PCR testing provider that occurred around October 2020. Initially, a positive case was identified by a defined Ct value of <40 in one of two PCR reactions using different primer pairs. Starting in October, a third PCR reaction was added to the protocol, with a positive case identified with a defined Ct value of <40 in at least two of the three reactions. This increased stringency may have helped exclude potential false positives, especially when the positive predictive value of the PCR-based test was low due to low prevalence in the population. These data could also potentially be explained by students having been infected during the summer and continuing to shed low levels of SARS-CoV-2 RNA in their nasopharynx at the time of the arrival tests in August. If this were the case, however, we would have expected the pre-arrival test that students took ahead of coming to campus to identify these individuals. Alternatively, students may have been infected during travel to campus, and therefore have been tested at a time prior to when they reached peak RNA levels. However, if this were the case, we would have expected that positive cases identified in the second round of arrival testing would have lower Ct values (higher RNA levels), which was not observed. Whatever the reason for this difference, it has implications for our wastewater results, as cases with higher Ct values (lower RNA levels) were significantly less likely to be associated with a positive wastewater result (Fig. 5D).

We observed no significant difference in the normalized SARS-CoV-2 RNA levels in wastewater based on the proximity of the wastewater sample to when a positive student case was identified (Fig. 5B), or based on the number of cases contributing to the waste stream (Fig. 5C). These results are contrary to other reports, in which wastewater SARS-CoV-2 RNA levels were observed to tightly correlate with case numbers (13, 14). We attribute this discrepancy to the relatively small number of people contributing to each waste stream in our study (hundreds vs. several thousand individuals in some municipal sewer sheds), and to the variable and sporadic nature of the flow rate at each sampling location. Composite samples from the lift stations tended to be more consistent than from the sewer access holes, where the flow rate changed dramatically on a minute-by-minute basis, and where the structure of the sewer caused pooling in some lines more than others. This caused marked differences in the amount and density of samples collected, and this variability could potentially lead to differences in the amount of biological material collected between the locations, or between samples from the same location. Additionally, during periods of inclement weather, some sampling locations could only be sampled with a single draw, rather than a 24-hour composite. These factors could therefore impact the relative concentration of SARS-CoV-2 RNA present in each sample, despite the use of RNase P RNA levels to normalize the SARS-CoV-2 RNA levels. Furthermore, the variable flow rate, as well as the lack of equipment to accurately measure this type of flow, made absolute quantification of SARS-CoV-2 genome copies per milliliter, (the standard used by several other studies; (11, 13)), impossible. We conclude that under these conditions, detection of SARS-CoV-2 RNA in the waste stream should be considered on a positive/negative basis, rather than attempting to use RNA quantification as a predictor of case load or timing.

One limitation of the study is that, due to the layout of campus infrastructure, several of the sample collection points did not only capture waste from student residences. In some cases, other campus buildings contributed to the waste stream (though every effort was made to minimize this), whereas in other cases, the residence halls also house additional administrative or classroom spaces. Additionally, custodial and facilities maintenance staff routinely accessed the buildings and may have contributed to the waste stream at the sampling locations. In one instance in the fall semester, a positive SARS-CoV-2 RNA signal in a wastewater sample was used to guide targeted PCR-based testing of students in residences, which did not identify any positive student cases. Concurrently, however, an employee that spent time in the space(s) contributing to that waste stream did test positive for SARS-CoV-2 infection, although whether they used restroom facilities in those location(s) is not known. Therefore, it is possible that other positive wastewater samples that did not coincide with a positive case may not be “false positives,” per se, but rather reflect other non-residents that contributed to the waste stream. It is also possible that these positive wastewater samples reflect infected students that were not identified by individual surveillance testing, either because they were not tested during their infection or their test results were falsely negative.

There are several additional limitations of this study that impact our ability to fully interpret the data. First, students that reported symptoms consistent with COVID-19 were tested using an in-house rapid nucleic acid amplification system that does not provide Ct values. Differences in viral dynamics at the time of testing (in surveillance tests vs. symptomatic testing) could skew our analysis of the Ct values of cases associated with positive wastewater detection of SARS-CoV-2. Second, the nature of our sampling strategy, with the collectors rotating between sampling locations on a two-to-three-day schedule, means that our samples were not evenly spaced, especially around times of arrival testing, when a majority of our positive cases were detected, and around times when students returned to residence halls following isolation. This confounds our analysis of wastewater RNA levels relative to the time of case detection or return, as it meant that some residences may have been sampled the morning after a student returned (potentially leading to a higher level of viral RNA shed into the waste stream), whereas others might not have been sampled until up to 72 h post-return, when shed RNA levels may have been lower. A sampling protocol that allowed for more continuous monitoring of wastewater RNA levels from residences following a return from isolation may have allowed for a more precise correlation between RNA levels and time from isolation return to be detected. This, however, would require more resources devoted to the project and would be beyond the scope of what our laboratory could handle. Finally, due to infrastructure architecture and geography, only around 73% of students lived in residences that could be easily monitored via the waste stream. This decreased the overall utility of wastewater surveillance as a means for managing emerging cases on campus.

Overall, based on the data presented here and our lived experience of managing the COVID-19 pandemic during the 2020-2021 academic year at Colgate University, we found that on a practical level, wastewater surveillance had more utility in the fall semester, when a smaller percentage of the community was subject to random PCR-based individual surveillance testing. The wastewater results could then be used to direct additional targeted testing to individual residences with positive wastewater RNA samples, and, with a positive predictive value of ∼54% for an isolated wastewater positive sample, this could be potentially be a more cost- and resource-effective means of detecting new positive cases than large-scale individual student testing. In the spring semester, with approximately half of the students in each residence were tested each week, by the time a positive wastewater sample was detected, it was more practical to wait for the individual surveillance results to become available than to target students in those residences for additional testing. This was further confounded when positive student cases returned to the residences following isolation, making it difficult to distinguish between a new infection and continued SARS-CoV-2 RNA shedding in previously-infected individuals. Additionally, the presence of cases in residences in which no wastewater SARS-CoV-2 RNA was detected suggests that wastewater surveillance of this type should not be used as the only means for the detection of cases on campus. As the price of personal surveillance testing, via pooled or individual samples, continues to decrease, we conclude that this should be viewed as a more effective means of controlling SARS-CoV-2 transmission on campus than wastewater testing of individual residence halls.

## Data Availability

All data related to this project is available at figshare.com, licensed CC by 4.0

https://doi.org/10.6084/m9.figshare.16563729.v1

## Authorship contribution statement

**G.H.H** initiated the project, developed the wastewater sampling strategy and SARS-CoV-2 RNA detection protocol, contributed to sample collection, analyzed and interpreted the data, and drafted the manuscript; **M.L**. contributed to sample collection, performed the RNA extraction and qPCR, analyzed the qPCR data, and edited the manuscript; **E.B**. contributed to sample collection; **E.L**. and **M.M** collected and assembled deidentified patient information, and edited the manuscript.

## Declaration of competing interest

The authors declare that they have no known competing financial interests or personal relationships that could have appeared to influence the work reported in this manuscript.

## Data availability statement

All data related to this project is available at figshare.com, licensed CC by 4.0

## Acknowledgments

The authors wish to thank David Larsen (Syracuse University) and Hyatt Green (SUNY ESF) for their initial input and advice on implementing a wastewater surveillance system. We also wish to thank Dan Gough, former Colgate University Associate Vice President for Campus Safety, and Environmental Health, and Emergency Management for assistance in initiating the project, and in procuring the wastewater sample collection devices. The authors are also grateful to Jim Albertina and the Colgate University Plumbing Department for their assistance in identifying, establishing, and maintaining wastewater sampling locations. We thank Karen Cheal, Neal Albert, and the office of Institutional Planning and Research for providing student housing data. We also thank Bineyam Taye and David Larsen for their thoughtful review of the manuscript. The authors are extremely grateful to the employees in Student Health Services, as well as the student and other volunteers who worked tirelessly in collecting student and employee patient samples throughout the year. Finally, the authors are extremely grateful to the Colgate University Health Analytics Team, especially Sev Flanigen, Tim Borfitz, and Mary Williams, for their work on behalf of the university, and for their thoughtful and productive analysis on the use of wastewater surveillance data to manage Colgate’s COVID-19 pandemic response. This project was funded entirely by Colgate University.

## Notes

### Competing Interest Statement

The authors have declared no competing interest.

### Author Declarations

This project was ruled as exempt from the review process by the Colgate University Institutional Review Board.

